# Assessing the prevalence of snake phobia among the general population in India

**DOI:** 10.1101/2024.06.12.24308826

**Authors:** Anika Salim, Gnaneswar Chandrasekharuni, José R Almeida, Rajendran Vaiyapuri, Harry F. Williams, Sundhararajan Arumugam, Subramanian Senthilkumaran, Ketan Patel, Timothy Williams, András Norbert Zsidó, Sakthivel Vaiyapuri

## Abstract

**Background:** A specific phobia is an anxiety disorder that is characterised by persistent and excessive fear in the presence of the object of the phobia. Animal phobias are the most prevalent forms of specific phobia among humans. Fear of snakes is present in non-human primates which suggests its evolutionary origins as the ability to detect the threat of snakes was critical for survival. Snake phobia is a critical factor in protecting snakes and mitigating snakebite burden. To date, only one standardised psychometric test [the Snake Questionnaire (SNAQ) developed in 1974] has been used to quantify snake phobia. Here, we estimated the level of snake phobia in India, where snakebites are highly prevalent using a modified version of the SNAQ (SNAQ12), which has previously demonstrated internal consistency, excellent reliability, and good discrimination between phobics and non-phobics in Hungary although it has never been tested among the general population in a snakebite-endemic country.

**Methodology/principal findings:** SNAQ12 was developed both in English and Tamil and validated by testing on several individuals. Then, the questionnaire was disseminated to members of the public through various methods including social media and in person through academic and clinical organisations. We received a total of 2032 responses, comprising 1086 [53.4%] males and 946 [46.6%] females.

**Conclusions/significance:** The results demonstrated good internal consistency in determining phobia amongst the population. The data suggests that males are more likely to be snake-phobic than females, in contrast to previous research that suggested that females are usually more snake-phobic. The use of the SNAQ12 allowed us to easily discriminate between individuals with phobia and non-clinical controls. This tool can be used as part of the One Health approach to better understand the relationships between snake phobia and snakebites and their impact on the mental health and well-being of vulnerable populations.

**Author Summary:** Snakebite envenoming often occurs due to unnecessary human-snake conflicts. Fear of snakes is a key factor in driving this conflict which frequently results in killing snakes. To protect humans from snakebites and snakes from humans, it is critical to estimate the level of snake phobia among vulnerable communities. Therefore, in this study, we used a robust online questionnaire (SNAQ12) to measure the level of snake phobia among members of the public in India, which is a snakebite-endemic country. SNAQ12 was developed in English and a local language, Tamil and validated before disseminating to members of the public through several methods. We received over 2000 responses, and the data analysis confirmed the internal consistency and robustness of this questionnaire for Indian communities. The data demonstrated that males are more phobic to snakes than females in India. The phobia was not dependent on any other factors such as age and education levels. While this study has some limitations, it forms the basis for further research to determine the level of phobia before and after snakebites and develop robust strategies to tackle this condition. We believe that preventing and treating snake phobia will aid in mitigating snakebite burden and snake conservation in India.

## Introduction

Snakebite envenoming (SBE) is a high-priority neglected tropical disease, that affects around 5.4 million people with approximately 150,000 deaths and 500,000 permanent disabilities every year worldwide [1-3]. India is the snakebite ‘capital’ of the world as it accounts for nearly half of the global burden of SBE-induced deaths, although the actual numbers are thought to be greatly underestimated due to the lack of reliable data [4-6]. Despite the high burden, very little is known about the fear of snakes among the general population living in rural areas in India as well as mental health sequelae caused by SBE in victims and their families [7]. A previous study highlighted the risk of Post Traumatic Stress Disorder (PTSD) in SBE patients and the necessity to establish better mental health safeguarding for them as this is often overlooked [7, 8]. Moreover, the relationships between snake phobias and their direct/indirect influences on SBE burden and snake conservation have not been analysed previously.

According to the World Health Organisation (ICD-11 WHO), phobias are anxiety disorders, evoked in people in situations that can be well-defined and pose no danger to the person with the phobia [9]. The spectrum of anxiety or fear-related disorders is classified into different subtypes. Specific phobia is a major subtype that is restricted to highly specific situations, for example, an extreme irrational fear of specific animals [9, 10]. Specific phobias are one of the most prevalent lifetime mental health disorders with prevalence rates ranging from 2.6% to 12.5% [11] and they cause long-term distress to many sufferers [9, 10, 12-14]. Snake phobia (ophidiophobia, a clinically relevant snake phobia) is a fear of snakes and is alleged to represent half of all animal phobias [15-17]. Indeed, our evolutionary origins may be responsible for this irrational fear as it might have provided our ancestors with an adaptive protective mechanism from venomous snakes for survival [18]. Hence, humans might have evolved to be predisposed to acquire a fear of snakes [19]. This has also been proposed in Seligman’s preparedness theory [20] which asserted that humans had acquired specific phobias due to inheriting a special sensitivity to stimuli (e.g., snakes) which represented a severe threat. Therefore, this evolutionary susceptibility to acquiring such an irrational fear can be easily induced in humans although it is resistant to treatment [20].

To date, very little is known about the prevalence of snake phobias among humans specifically in snakebite-endemic countries, where the risks of SBE-induced deaths, disabilities and socioeconomic impacts are high. Snake phobia results in poor management of the human-snake conflict and mental health prognosis in SBE victims. Therefore, it is imperative to estimate snake phobia among vulnerable populations and develop appropriate initiatives to reduce such fear of snakes to save snakes as they play an important part in our ecosystem and mitigate the SBE burden. Despite its high prevalence, only one test to measure snake phobia was developed by Klorman and colleagues in 1974 [21]. This was then revised by Zsido et al. 2018 [22] to create the SNAQ12, which is a shorter psychometric test for use in clinical settings and this has been proven to be effective, economical, and efficient in Hungary. However, the SNAQ12 has not been tested extensively in wider populations in other countries, specifically where snakebites are prevalent. Here, we validated and used the SNAQ12 in India (with a specific focus on Tamil Nadu), where the SBE burden is high.

## Materials and Methods

### Translation and validation of the SNAQ12

The original SNAQ [21] is a 30-item self-reported measure of fear and phobia of snakes. A shorter version of this survey, the SNAQ12 was developed and validated by researchers in the Hungarian language and was shown to have excellent psychometric properties (Zsido et al. 2018). The SNAQ-12 is a 12-item questionnaire employing a discriminatory scale, where participants indicate whether or not they agree with a statement. This test has excellent discriminatory power, hence it is useful as a diagnostic tool for snake phobia. Individuals with scores of ≥8 on the SNAQ-12 are considered to be phobic to snakes. Therefore, the SNAQ12 can be recommended for use in clinical practice for fast and accurate estimations of a respondent’s fear of snakes. The copyright author provided written consent for the SNAQ12 to be used in this study and to be translated into Tamil. The SNAQ12 was adapted and translated into the Tamil language by lead authors who are fluent in both Tamil and English. The validation of the SNAQ12 was then carried out by our colleagues in India by collecting responses and feedback from 100 people. Based on their feedback, an expert panel of researchers ensured the translation was as close as possible to the original SNAQ12 and accessible to both genders and ages. The online survey incorporated both the English and Tamil texts for all participants (**Supplementary information**).

### Patient and public involvement statement

The patients and members of the public were not directly involved in the study design, data collection, analysis and writing of the manuscript mainly due to their limited availability to attend several meetings that are often arranged during day times when they are also occupied with their daily work. However, all patients provided written consent to collect these data and to publish them in scientific journals. Involvement in this study was completely voluntary and participants could withdraw their consent at any time during this study. The participation did not result in any implications for their treatment and outcomes. We will ensure that the results of this study are disseminated to study participants and wider communities through scientific publications, which might be followed by press releases in media in the local language and English.

### Study design and data collection

This study was performed according to the Declaration of Helsinki and approved by the Institutional Ethics Committee of Toxiven Biotech Private Limited (reference: ICMR-Toxiven Ethics 2022/001) and the University of Reading Research Ethics Committee (reference: UREC 23/05). Written informed consent was obtained from all study participants involved in the study to anonymously analyse and publish the data. This prospective study was conducted between September 2022 and February 2023, and we aimed to recruit at least 2000 participants across Tamil Nadu. Participants living in other Indian states were also allowed to participate following exclusion (anyone aged less than 18 and inability to read and provide consent) and inclusion (anyone aged over 18 with the ability to read and provide consent) criteria. Adults over the age of 18 were invited, and there were no other inclusion or exclusion criteria. Participants were not compensated as participation was completely voluntary. The survey was administered online using Jisc Online Surveys (JISC 2020) and respondents were recruited through social media and our academic and social networks in India. The link to the survey with a short and informative description of the study was posted on various forums, social networks, and mailing lists to ensure maximal participation. To ensure participation from a variety of demographic, socioeconomic and educational backgrounds, invitations to the survey were posted on various forums and mailing lists. Participants were also directly approached in several colleges in Tamil Nadu.

### Statistical methods

All statistical analyses were performed using SPSS (version 26) (IBM, UK), JASP (version 0.17.2) (JASP team 2023, Amsterdam, The Netherlands), GraphPad Prism (version 8.0.0) (GraphPad Prism Inc, USA) and Jamovi (version 2.3) (www.jamovi.org).

## Results

### Study population

The SNAQ12 questions were available in both English and Tamil and were developed as an online survey which can be completed via any device with internet access. Following validation, the questionnaire was circulated to members of the public and students in colleges and universities via multiple routes (more details are provided in the methods section). A total of 2032 responses were received across a wide geographical distribution of Tamil Nadu (a major state in India with a high SBE burden) as well as other parts of India (**Figure 1A**). Tamil Nadu is comprised of 38 districts, and we received responses from all districts confirming the representative nature of the data. The following districts in Tamil Nadu displayed the highest responses: Coimbatore [689 (33.9%)], Chennai (the capital city of Tamil Nadu) [180 (8.9%)], Tiruchirappalli [123 (6.1%)] and Salem [89 (4.4%)]. A total of 256 responses were received from other Indian states.

**Figure 1:**
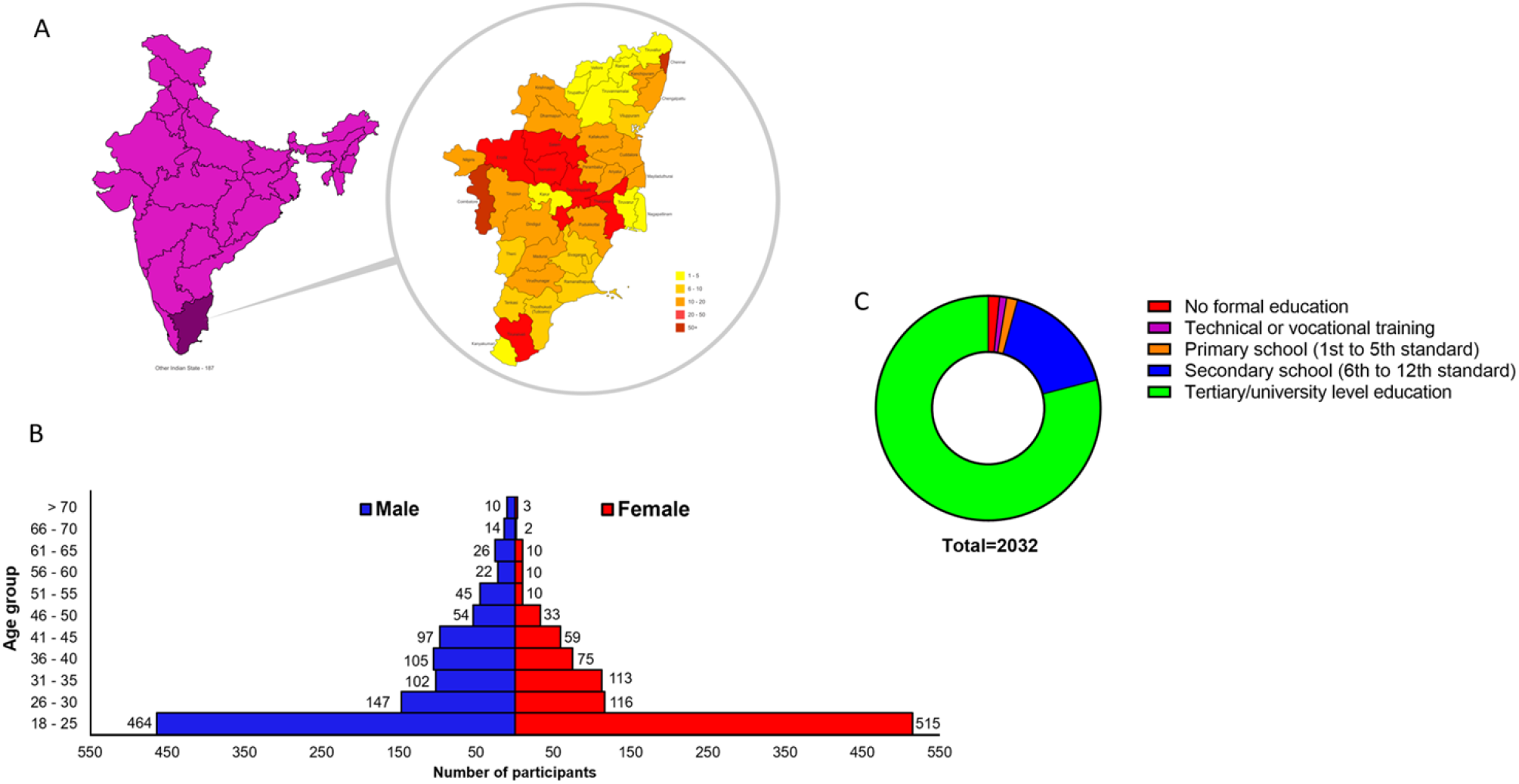
Characteristics of the study population. **A**) a map of India showing the location of Tamil Nadu and the geographical heatmap (insert) displaying participant responses from the 38 districts in Tamil Nadu, India. Districts are colour-graded according to the number of responses received. **B**) the total number of participants in this study was organised based on their gender and age groups. **C**) the distribution of participants based on their level of education.

Of the 2032 participants, 1086 (53.4%) were males and 946 (46.6%) were females. Moreover, 979 (48.2%) [464 (22.8%) males and 515 (25.3%) females] participants were in the age group of 18 to 25 years old, 263 (12.9%) [147 (7.2%) males and 116 (5.7%) females] were between 26 and 30 years, 215 (10.5%) [102 (5%) males and 113 (5.5%) females] were between 31 and 35 years. 180 (8.8%) [105 (5.16%) males and 75 (3.69%) females] were between 36 and 40 years, 156 (7.6%) [97 (4.77%) males and 59 (2.9%) females] were between 41 and 45 years, 87 (4.2%) [54 (2.65%) males and 33 (1.62%) females] were between 46 and 50 years, 55 (2.7%) [45 (2.21%) males and 10 (0.49%) females] were between the ages of 51 and 55, 32 (1.5%) [22 (1.08%) males and 10 (0.49%) females] were between the ages of 56 and 60, 36 (1.7%) [26 (1.27%) males and 10 (0.49%) females] were between the ages of 61 and 65, 16 (0.7%) [14 (0.68%) males and 2 (0.09%) females] between the ages of 66 and 70 and 13 (0.6%) [10 (0.49%) males and 3 (0.14%) females] participants were over the age of 70 (**Figure 1B**). When the age groups were ranked with the number of participants, the median age of female participants was between 26 and 30 years and males was between 31 and 35 years.

Of the 2032 participants, 1606 (79%) had tertiary university level education, 340 (16.7%) had secondary school education, 32 (1.6%) had primary school education, 21 (1%) had technical/vocational training and 33 (1.6%) had no formal education (**Figure 1C**).

### Confirmatory factor analysis ascertains the reliability of the SNAQ12

To determine the reliability of the SNAQ12 among this study population, a confirmatory factor analysis was performed using the diagonally weighted least squares (DWLS) estimator. For model fit, the comparative fit index (CFI), the Tucker-Lewis index (TLI), the root mean square error of approximation (RMSEA), and the standardized root mean squared residual index (SRMR) were used. They yielded an adequate level of fit on this sample (CFI = .992, TLI = .989, RMSEA = .057 95% CI = .051 to .064, and SRMR = .057). The factor loadings varied between .63 and .84. These results indicate that the one-factor solution fits the data and the SNAQ12 is a reliable tool in this study population to measure fear of snakes.

Moreover, to demonstrate the internal consistency of the SNAQ12 among this study population, Cronbach’s alpha which calculates the pairwise correlations between items in a survey was used and it measured 0.84 (0.8 ≤ α < 0.9). Similarly, the McDonald’s omega score was measured as 0.84. This illustrates that the SNAQ12 has good internal consistency in measuring snake phobia among this study population in India.

### Males are more snake-phobic than females

The responses from all study participants for different questions are shown in **Figure 2**. The overall average score was 7.5 for all 2032 participants with a median score of 8 (SD - 3.26). 1061 (52.2%) participants were identified as having a potential snake phobia as they scored 8 or above on the SNAQ12 with an average score of 10 and a median score of 10 (SD - 1.5). A total of 971 (47.8%) participants were identified as having no snake phobia as they scored less than 8, with the average score across this population of 4.6, with a median score of 5 (SD - 2.1).

**Figure 2:**
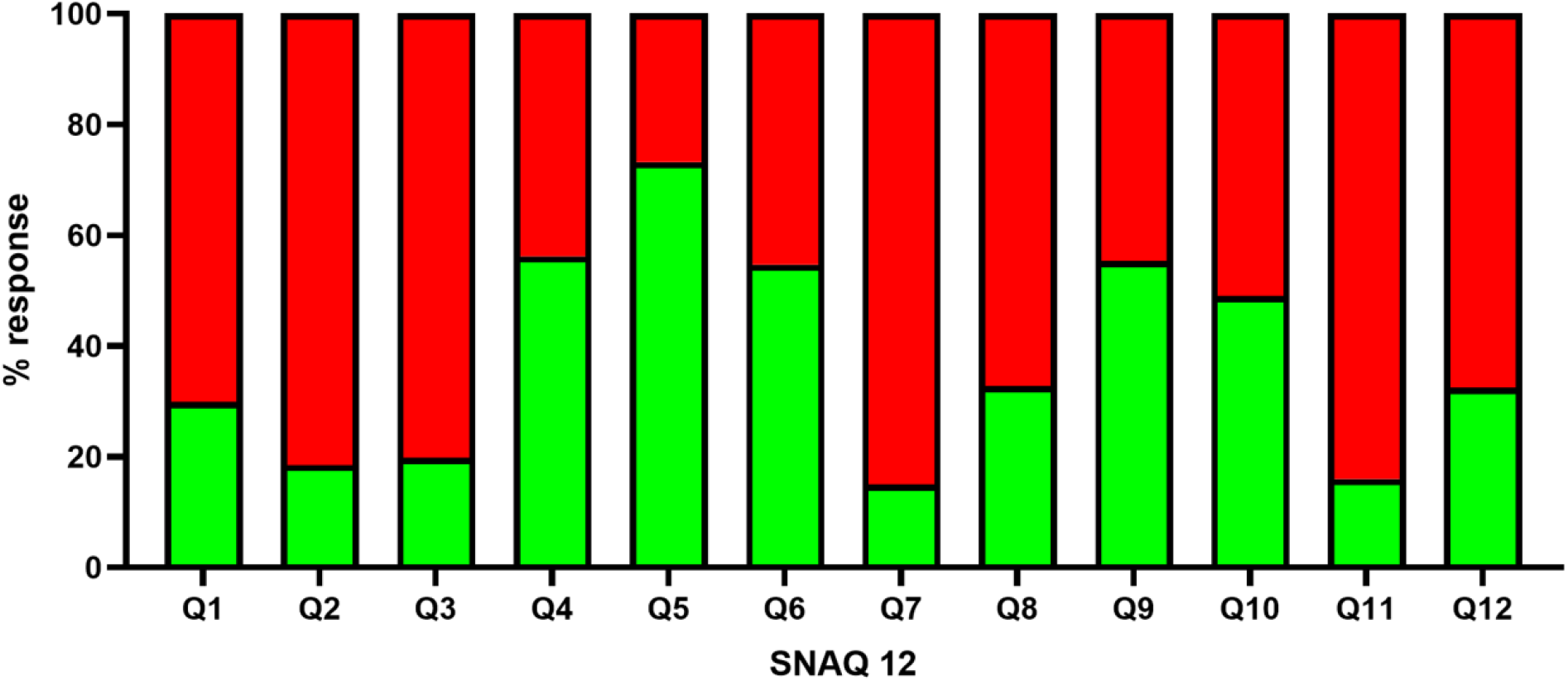
Total responses for individual questions in SNAQ12. The total percentage of responses from participants for each SNAQ12 question is shown. Green indicates the percentage of participants that answered ‘yes’ and red indicates the percentage of participants that answered ‘no’ to each question. The questions used in this study are shown in supplementary information.

Furthermore, the average score for the female population (946) was 6.4 with a median of 6 (SD - 3.3). Among them, 359 (38% of the total 946) females were considered to have a phobia as they scored an average of 9.8, a median of 10 (SD - 1.4) and 587 (62% of the total 946) females were considered to have no phobia with an average score of 4.3, a median of 5 (SD - 2.1) (**Table 1**). Similarly, 1086 participants were males, and their average score was 8.4, a median of 9 (SD - 3). 702 (64.6% of the total 1086) males were considered to have a snake phobia and their average score was 10.2 with a median of 10 (SD - 1.4). However, 384 (35.4% of the total 1086) males were considered to have no phobia as they had an average score of 5 with a median of 6 (SD - 1.9) (**Table 1**). Notably, there is a significant difference in the average scores for both males and females (p=<0.0001; t=14.231; df=2030) suggesting that males (∼65%) are more snake-phobic than females (38%).

**Table 1:** Mean and median SNAQ scores of males and females in the study population. This highlights the number of phobic and non-phobic individuals as well as the mean and median age groups of males and females.

|  | **Mean score** | **Median score** | **STDEV** | **Phobia ≥8** | **No Phobia <8** |
| --- | --- | --- | --- | --- | --- |
| Total number (2032) | 7.5 | 8 | 3.3 | 1061 | 971 |
| Mean and median age group: **26-30** | 7.9 | 8 | 3.3 | 146 | 117 |
| Total number of females (946) | 6.4 | 6 | 3.3 | 359 | 587 |
| Median age group of females: **18-25** | 6.3 | 6 | 3.2 | 190 | 325 |
| Mean age group of females: **26-30** | 7 | 7 | 3.4 | 51 | 65 |
| Total number of males (1086) | 8.4 | 9 | 3 | 702 | 384 |
| Median age group of males: **26 - 30** | 8.6 | 9 | 3.1 | 95 | 52 |
| Mean age group of males: **31 - 35** | 8.3 | 9 | 3.1 | 65 | 37 |

### Snake phobia is not dependent on age group

We then analysed the presence of snake phobia among different age groups (**Table 2**). The results demonstrate that 495 of 18-25 (50.6% of a total 979), 146 of 26-30 (55.5% of 263), 107 of 31-35 (49.8% of 215), 97 of 36-40 (50.6% of 180), 82 of 41-45 (52.6% of 156), 45 of 46-50 (51.7% of 87), 29 of 51-55 (52.7% of 55), 15 of 56-60 (47% of 32), 27 of 61-65 (75% of 36), 12 of 66-70 (75% of 16), and 6 of >70 (46% of 13) were identified as phobic for snakes (**Figure 3**). Although the percentage of people who are phobic of snakes in the age range of 61-70 is high, there was no significant difference between the average scores across the age groups. These data suggest that age does not impact the presence of snake phobia in this study population. A correlation analysis was also conducted and found that there was no significant correlation between the age groups amongst the total population and average SNAQ scores [*r*(2032)=.01, P=.53]. However, when comparing the phobias between males (**Figure 3A**) and females (**Figure 3B**) among different age groups, males are significantly more phobic than females in all age groups. The correlation analysis found that there was no significant correlation between the different age groups within the male population [*r*(1084) = -.05, P = .07 and in females *r*(944) = -.01, P = .64].

**Table 2:** Mean and median SNAQ scores of different age groups in the study population. This highlights the number of phobic and non-phobic individuals in different age groups.

| **Group** | **N** | **Mean** | **Median** | **SD** |
| --- | --- | --- | --- | --- |
| Total | 2032 | 7.5 | 8 | 3.3 |
| Total non phobic | 971 | 4.6 | 5 | 2.1 |
| Total phobic | 1061 | 10 | 10 | 1.5 |
| **Age groups** | **N** | **Mean** | **Median** | **SD** |
| **18 - 25** | **979** | **7.4** | **8** | **3.3** |
| 18-25 non phobic | 484 | 4.6 | 5 | 2.1 |
| 18-25 phobic | 495 | 10 | 10 | 1.5 |
| **26 - 30** | **263** | **7.9** | **8** | **3.3** |
| 26 - 30 non phobic | 117 | 4.7 | 5 | 2.1 |
| 26 - 30 phobic | 146 | 10.4 | 11 | 1.4 |
| **31 - 35** | **215** | **7.3** | **7** | **3.4** |
| 31 - 35 non phobic | 108 | 4.4 | 5 | 2.2 |
| 31 - 35 phobic | 107 | 10.2 | 11 | 1.5 |
| **36 - 40** | **180** | **7.4** | **8** | **3.2** |
| 36 - 40 non phobic | 83 | 4.6 | 5 | 2 |
| 36 - 40 phobic | 97 | 9.8 | 9 | 1.5 |
| **41 - 45** | **156** | **7.4** | **8** | **3.1** |
| 41 - 45 non phobic | 74 | 4.7 | 5 | 2.1 |
| 41 - 45 phobic | 82 | 9.9 | 10 | 1.3 |
| **46 - 50** | **87** | **7.1** | **8** | **3.4** |
| 46 - 50 non phobic | 42 | 4.2 | 5 | 2.4 |
| 46 - 50 phobic | 45 | 9.8 | 10 | 1.5 |
| **51 - 55** | **55** | **7.5** | **8** | **3.2** |
| 51 - 55 non phobic | 26 | 4.7 | 5 | 2.2 |
| 51 - 55 phobic | 29 | 10.1 | 10 | 1.2 |
| **56 - 60** | **32** | **7** | **7** | **3.5** |
| 56 - 60 non phobic | 17 | 4.4 | 3 | 2.3 |
| 56 - 60 phobic | 15 | 10.1 | 10 | 1.6 |
| 61 - 65 | **36** | **8.7** | **9** | **2.4** |
| 61 - 65 non phobic | 9 | 5.3 | 6 | 1.6 |
| 61 - 65 phobic | 27 | 9.8 | 10 | 1.4 |
| **66 - 70** | **16** | **8.4** | **9** | **3.4** |
| 66 - 70 non phobic | 4 | 3.8 | 4 | 2.9 |
| 66 - 70 phobic | 12 | 9.9 | 9.5 | 1.6 |
| **> 70** | **13** | **7.1** | **7** | **2.5** |
| > 70 non phobic | 7 | 5.3 | 5 | 1.2 |
| > 70 phobic | 6 | 9.3 | 9 | 1.5 |

**Figure 3:**
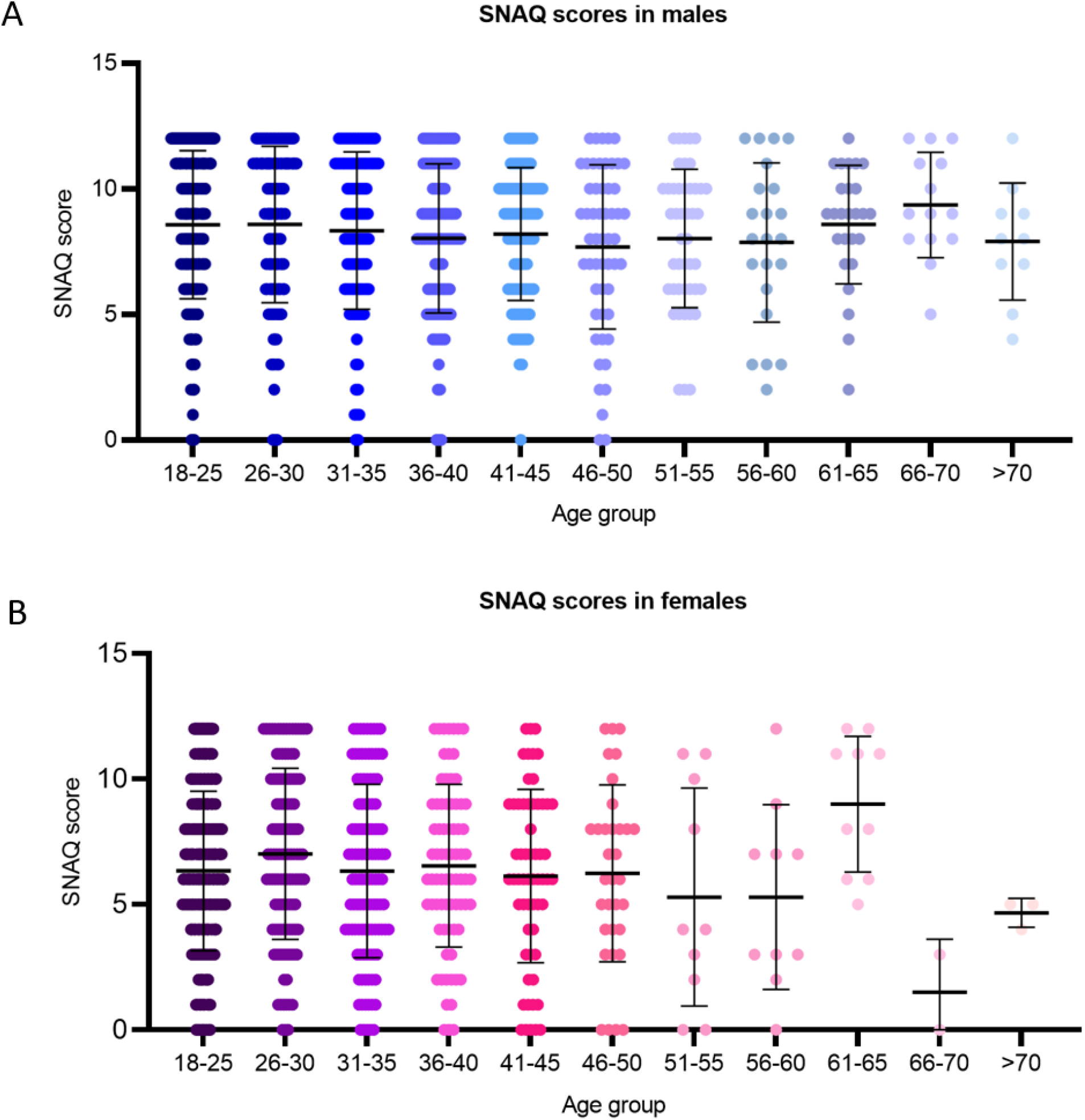
The distribution of SNAQ12 scores in males (**A**) and females (**B**) across different age groups.

### The level of education does not impact snake phobia

We then analysed the impact of education on developing snake phobia among the study participants. 843 (52.5% of the total of 1606) people who received university-level education were found to have snake phobia [average score - 10.1; median - 10; SD - 1.5]. Similarly, 179 (52.6% of 340) who received secondary school level education were snake phobic [average score - 9.9; median - 10; SD - 1.4]. Moreover, 16 (50%) [average score - 10.3; median - 10; SD - 1.6] who received primary school education, 10 (47,6%) [average score - 9.6; median - 9.5; SD - 1.6] who received technical/vocational training and 13 (39.4%) [average score - 10.2; median - 10; SD - 1.5] who have not received any formal education were found to have a snake phobia. There was no significant difference between the average SNAQ scores across the various educational groups. This suggests that the level of education does not impact the presence of snake phobia in this study population.

## Discussion

SBE is a predominant occupational health hazard in rural, impoverished, agricultural communities in developing countries [6, 23]. The fear of snakes that develops naturally among rural populations persists throughout their lifetime, and it often affects their mental health and well-being. Although most people are aware that not all snakes are venomous or dangerous, their fear forces them to kill any snake that they see in their dwellings as well as in their natural habitats [24]. This has immense implications for human-snake conflicts and reptile conservation, particularly in vulnerable and diverse biomes, as snakes play a vital predatory role in our ecosystems and food webs albeit not least in balancing rodent populations in agricultural communities. This in turn may reduce the population of snakes and ultimately increase the population of rodents which can affect crop production [25]. Moreover, the fear of snakes often leads to increasing human-snake conflicts, SBE burden, and perpetuation of the cycle of poverty due to long-term complications that are deeply associated with economic and social ramifications [26]. For example, some people in rural communities are reluctant to improve their awareness about snakes and SBE mainly because of their phobia of snakes [27]. Thus, the global SBE crisis is a potentially lethal and debilitating consequence derived from a network of multiple interconnected factors that exacerbate the vulnerability of rural communities in low-resource regions [28]. In this multidimensional scenario, understanding the overlap and domino effect between human activities, distribution and ecology of snakes, fear, human-wildlife interactions, and snakebites should be a fundamental piece of the global plan of action against this neglected disease. Additionally, the relationship between these key factors is complex, highly dynamic, and context-dependent, which must take into account the religious and cultural aspects of the most affected populations. Mythical and religious beliefs play a key role in the development of snake phobia and consequently, in SBE burden [29]. As a result, synergistic actions, and efforts, such as conflict prevention strategies, community-based programs, sustainable coexistence promotion techniques, snake conservation initiatives and multilevel investigations of the different factors or priority elements mentioned above are crucial to minimise the huge impact of this challenge [30]. Understanding the scope of snake phobias in snakebite endemic communities allows us to develop targeted educational strategies to improve community understanding and implement productive approaches to snake conservation that not only promote positive wildlife interactions but also minimise snakebite incidents. Therefore, it is critical to tackle this issue by estimating snake phobia and developing better strategies to reduce the fear among vulnerable populations, enhance the conservation of snakes and improve the clinical management of SBE.

The data from this study emphasises that males are more likely to be snake-phobic across all age groups compared to females. The age and level of education appeared to not impact the development of snake phobia. This is contrary to most previous studies where the female members of a population were usually reported to be the most phobic of snakes [14, 16, 31]. When it comes to the epidemiology of anxiety disorders, they are often significantly more prevalent in females compared to males. It is widely accepted that common fears have a higher incidence in childhood and then they rapidly taper off during adolescence and post-puberty [31]. However, our data highlight that the presence of a snake phobia does not decrease with age, in either males or females. When comparing both genders and their respective age groups, males proved to be more significantly phobic compared to females across all ages. It has been reported that the presence of sex differences in specific phobias is more apparent after puberty and the ratings are usually lower when age is increased [31]. The developmental process of adolescent fear responsiveness and acquisition has not been greatly characterised [32]. Unlike other anxiety disorders, it is generally assumed that phobias can be acquired at any time, from childhood fear acquisition to adolescence, and adulthood. Therefore, age is not a restriction to developing, maintaining, and eliminating the fear of snakes as demonstrated in this study. Similarly, many participants in this study received university-level education, but still, most of them were found to be phobic of snakes. This suggests that education is not a factor in developing or removing phobia, and the current education may not help alleviate the fear of snakes among the general population. A more specific curriculum to alleviate the fear of snakes would aid in reducing snake phobia among students, and they may continue to live without the fear for the rest of their lives. A gentle harmless exposure of snakes to phobic people is also likely to reduce their fear of snakes [24]. In a specific population of Nigeria, the increased exposure to snakes has proven to have reduced phobia among the communities although a single snakebite resulted in the opposite effect and killing of snakes [24]. Similarly, a guided internet-delivered exposure strategy has been proposed and evaluated as a promising solution [33, 34]. The findings of this study support its potential and applicability to reduce snake phobia. In a clinical setting, other management strategies may also be useful, such as cognitive behavioural interventions and pharmacotherapy.

Many theories have been put forward for why humans develop or are predisposed to specific snake phobias including the conditioning theory of fear acquisition, social learning theory and the preparedness theory [35-37]. Familial transmission or increased frequency of social or indirect adverse exposure could account for why males in this study population have a higher incidence of phobia. For example, witnessing a reaction to a snake by someone suffering from snake phobia and higher exposure to snakes due to increased presence in agricultural activities could explain the higher phobic incidence among males. Fear of snakes has long been associated with evolutionary origins in protecting and ensuring our survival. The snake detection theory even suggests that the evolution of the primate visual system was innately adapted to detect threats such as snakes better in the environment [38]. Therefore, the ability of our ancestors to identify a snake and potential risk at any age would have been critical to early human survival. This could explain why there was no significance in snake phobia across the age groups in this study. Behavioural and electrophysiological research and the use of event-related potentials and earlier posterior negativity amplitude tests with brain imaging over the past decade have aided in improving our understanding of snake phobias. These studies have illustrated how snakes are not only easily identified by humans but also by primates held in captivity who have never been exposed to a real snake. This provides evidence of the fact that humans are naturally predisposed to rapidly identify a snake [38, 39].

Several studies have highlighted that there is a close association between specific phobias and other psychiatric and non-psychiatric disorders [40]. Indeed, specific phobias can result in co-morbidities such as cardiovascular disease, migraine and thyroid disorders [22]. SBE can be considered a traumatic episode for a patient and developing/having a snake phobia can worsen this situation. Therefore, effective, and economical psychometric tools that have been appropriately validated among vulnerable populations are vital for healthcare providers and mental health professionals when working with SBE patients to provide better care. The clinical assessments of snake phobia have mainly been conducted in developed countries where SBE is not a major concern. For the first time, in this study, the SNAQ12 has been used on the general population of India where SBE is an endemic medical issue. The SNAQ12 is a modified version of the original SNAQ with only 12 questions and the reliability and efficiency of this tool have been established previously in Hungary [22]. We developed the SNAQ12 in a bilingual format which can be easily accessed and completed online using any device. The ease of use, lack of interpretability issues and low economic footprint mean the SNAQ12 can be a useful assessment tool for researchers, clinicians, and caregivers to better understand the level of snake phobia in patients and improve treatment outcomes. This will also improve the treatment-seeking behaviour and reduce the long-term impacts on SBE patients as well as others.

Our perceptions and interactions with the environment influence our relationships with the communities. Human-animal conflict is more common now than ever before and negative emotions and experiences are thought to underpin the maintenance and longevity of specific phobias including snake phobia [41]. Our intrinsic negative attitudes to snakes resulting in humans attacking and killing them have been one of the causes of the great losses to our reptile biodiversity. Killing snakes has a downstream ecological impact on the environment. Due to the threat that snakes pose in communities where SBE is common, they are more likely to be killed [42]. Therefore, it is critical to conserve snakes to promote their support in agriculture while saving lives from snakes. The deployment of the SNAQ12 in assessing the attitudes of populations to snakes and subsequent actions may also aid in reducing the killing of snakes by vulnerable communities. Better education about the importance of snakes among children and adolescent students in schools and colleges/universities will further promote the cohabitation of humans and snakes. Overall, this study provides an overview of the status of snake phobia in India, which is considered the capital of SBE due to the high number of deaths. Using the SNAQ12 in other parts of India as well as other countries will further establish the relationships between snake phobia and SBE burden.

### Limitations

Despite obtaining a high response rate, this study has several limitations. The SNAQ12 was completed online using electronic devices and therefore, this might have prevented people who do not have access to such devices from completing the survey. Moreover, some people might have felt uncomfortable completing a study that is related to snakes. Similarly, this survey was not possible to complete without an assistant for people who were unable to read Tamil or English. Due to ethical concerns, this study only collected responses from people who were aged 18 and older. As specific phobias are some of the most prevalent anxiety disorders in children, this should be considered in future studies along with surveying adolescents to estimate the level of phobia in under-18s/non-adults. In addition, it would be better to include an equal number of males and females in different age groups to quantify their overall SNAQ scores. Future research is also required to establish the relationship between snake phobia and snakebite burden among vulnerable populations.

## Data Availability

All data are included within this article

## Acknowledgements

We would like to thank all the study participants for their time and for completing the SNAQ12 in this study. We also extend our thanks to several collaborators, snake rescuers, NGOs and volunteers for disseminating this survey among the students and members of the public. We are grateful to the Medical Research Council, UK (Grant reference: MR/W019353/1 and ITTP PhD Studentship) for their funding support.

## References

1. Kasturiratne A, Wickremasinghe AR, de Silva N, Gunawardena NK, Pathmeswaran A, Premaratna R, et al. The global burden of snakebite: a literature analysis and modelling based on regional estimates of envenoming and deaths. PLoS Med. 2008;5(11):e218. doi: 10.1371/journal.pmed.0050218. PubMed PMID: 18986210. PubMed PMID: PMCPMC2577696.

2. Longbottom J, Shearer FM, Devine M, Alcoba G, Chappuis F, Weiss DJ, et al. Vulnerability to snakebite envenoming: a global mapping of hotspots. The Lancet. 2018;392(10148):673–84. doi: 10.1016/S0140-6736(18)31224-8.

3. Gutiérrez JM, Calvete JJ, Habib AG, Harrison RA, Williams DJ, Warrell DA. Snakebite envenoming. Nat Rev Dis Primers. 2017;3:17063. Epub 20170914. doi: 10.1038/nrdp.2017.63. PubMed PMID: 28905944.

4. Suraweera W, Warrell D, Whitaker R, Menon G, Rodrigues R, Fu SH, et al. Trends in snakebite deaths in India from 2000 to 2019 in a nationally representative mortality study. eLife. 2020;9:e54076. doi: 10.7554/eLife.54076.

5. Mohapatra B, Warrell DA, Suraweera W, Bhatia P, Dhingra N, Jotkar RM, et al. Snakebite Mortality in India: A Nationally Representative Mortality Survey. PLOS Neglected Tropical Diseases. 2011;5(4):e1018. doi: 10.1371/journal.pntd.0001018.

6. Vaiyapuri S, Vaiyapuri R, Ashokan R, Ramasamy K, Nattamaisundar K, Jeyaraj A, et al. Snakebite and its socio-economic impact on the rural population of Tamil Nadu, India. PLoS One. 2013;8(11):e80090. Epub 20131121. doi: 10.1371/journal.pone.0080090. PubMed PMID: 24278244. PubMed PMID: PMCPMC3836953.

7. Millán-González R, Monge-Morales LF, De La Cruz-Villalobos N, Bonilla-Murillo F, Gutiérrez JM. Bothrops asper bite and post-traumatic stress disorder in Costa Rica: Report of two cases. Toxicon. 2023;231:107199. Epub 20230614. doi: 10.1016/j.toxicon.2023.107199. PubMed PMID: 37328114.

8. Habib ZG, Salihu AS, Hamza M, Yakasai AM, Iliyasu G, Yola IM, et al. Posttraumatic stress disorder and psycho-social impairment following snakebite in Northeastern Nigeria. Int J Psychiatry Med. 2021;56(2):97-115. Epub 20200326. doi: 10.1177/0091217420913400. PubMed PMID: 32216497.

9. Eaton WW, Bienvenu OJ, Miloyan B. Specific phobias. Lancet Psychiatry. 2018;5(8):678–86. doi: 10.1016/s2215-0366(18)30169-x. PubMed PMID: 30060873. PubMed PMID: PMCPMC7233312.

10. LeBeau RT, Glenn D, Liao B, Wittchen H-U, Beesdo-Baum K, Ollendick T, et al. Specific phobia: a review of DSM-IV specific phobia and preliminary recommendations for DSM-V. Depression and Anxiety. 2010;27(2):148–67. doi: 10.1002/da.20655.

11. Wardenaar KJ, Lim CCW, Al-Hamzawi AO, Alonso J, Andrade LH, Benjet C, et al. The cross-national epidemiology of specific phobia in the World Mental Health Surveys. Psychological Medicine. 2017;47(10):1744-60. Epub 2017/02/22. doi: 10.1017/S0033291717000174.

12. Becker ES, Rinck M, Türke V, Kause P, Goodwin R, Neumer S, et al. Epidemiology of specific phobia subtypes: findings from the Dresden Mental Health Study. Eur Psychiatry. 2007;22(2):69-74. Epub 20061208. doi: 10.1016/j.eurpsy.2006.09.006. PubMed PMID: 17157482.

13. Grenier S, Schuurmans J, Goldfarb M, Préville M, Boyer R, O’Connor K, et al. The epidemiology of specific phobia and subthreshold fear subtypes in a community-based sample of older adults. Depress Anxiety. 2011;28(6):456-63. Epub 20110311. doi: 10.1002/da.20812. PubMed PMID: 21400642.

14. Fredrikson M, Annas P, Fischer H, Wik G. Gender and age differences in the prevalence of specific fears and phobias. Behaviour Research and Therapy. 1996;34(1):33–9. doi: 10.1016/0005-7967(95)00048-3.

15. Klieger DM. The Snake Anxiety Questionnaire as a Measure of Ophidophobia. Educational and Psychological Measurement. 1987;47(2):449–59. doi: 10.1177/0013164487472017.

16. Polák J, Sedláčková K, Nácar D, Landová E, Frynta D. Fear the serpent: A psychometric study of snake phobia. Psychiatry Res. 2016;242:163-8. Epub 20160531. doi: 10.1016/j.psychres.2016.05.024. PubMed PMID: 27280527.

17. Davey GCL. Self-reported fears to common indigenous animals in an adult UK population: The role of disgust sensitivity. British Journal of Psychology. 1994;85(4):541–54. doi: 10.1111/j.2044-8295.1994.tb02540.x.

18. Weiss L, Brandl P, Frynta D. Fear reactions to snakes in naïve mouse lemurs and pig-tailed macaques. Primates. 2015;56(3):279–84. doi: 10.1007/s10329-015-0473-3.

19. LoBue V, DeLoache JS. Detecting the Snake in the Grass:Attention to Fear-Relevant Stimuli by Adults and Young Children. Psychological Science. 2008;19(3):284–9. doi: 10.1111/j.1467-9280.2008.02081.x. PubMed PMID: 18315802.

20. Seligman MEP. Phobias and preparedness. Behavior Therapy. 1971;2(3):307–20. doi: 10.1016/S0005-7894(71)80064-3.

21. Klorman R, Weerts TC, Hastings JE, Melamed BG, Lang PJ. Psychometric description of some specific-fear questionnaires. Behavior Therapy. 1974;5(3):401–9. doi: 10.1016/S0005-7894(74)80008-0.

22. Zsido AN, Arato N, Inhof O, Janszky J, Darnai G. Short versions of two specific phobia measures: The snake and the spider questionnaires. Journal of Anxiety Disorders. 2018;54:11–6. doi: 10.1016/j.janxdis.2017.12.002.

23. Harrison RA, Hargreaves A, Wagstaff SC, Faragher B, Lalloo DG. Snake envenoming: a disease of poverty. PLoS Negl Trop Dis. 2009;3(12):e569. Epub 20091222. doi: 10.1371/journal.pntd.0000569. PubMed PMID: 20027216. PubMed PMID: PMCPMC2791200.

24. Coelho CM, Polák J, Suttiwan P, Zsido AN. Fear inoculation among snake experts. BMC Psychiatry. 2021;21(1):539. doi: 10.1186/s12888-021-03553-z.

25. Kontsiotis VJ, Rapti A, Liordos V. Public attitudes towards venomous and non-venomous snakes. Science of The Total Environment. 2022;831:154918. doi: 10.1016/j.scitotenv.2022.154918.

26. Landry Yuan F, Devan-Song A, Yue S, Bonebrake TC. Snakebite Management and One Health in Asia Using an Integrated Historical, Social, And Ecological Framework. Am J Trop Med Hyg. 2021;106(2):384-8. Epub 20211206. doi: 10.4269/ajtmh.21-0848. PubMed PMID: 34872063. PubMed PMID: PMCPMC8832943.

27. Vaiyapuri S, Kadam P, Chandrasekharuni G, Oliveira IS, Senthilkumaran S, Salim A, et al. Multifaceted community health education programs as powerful tools to mitigate snakebite-induced deaths, disabilities, and socioeconomic burden. Toxicon: X. 2023;17:100147. doi: 10.1016/j.toxcx.2022.100147.

28. Warrell DA, Williams DJ. Clinical aspects of snakebite envenoming and its treatment in low-resource settings. Lancet. 2023;401(10385):1382-98. Epub 20230314. doi: 10.1016/s0140-6736(23)00002-8. PubMed PMID: 36931290.

29. Senthilkumaran S, Arathisenthil SV, Williams J, Williams HF, Thirumalaikolundusubramanian P, Patel K, et al. An Unnecessary Russell’s Viper Bite on the Tongue Due to Live Snake Worship and Dangerous First Aid Emphasise the Urgent Need for Stringent Policies. Toxins (Basel). 2022;14(12). Epub 20221122. doi: 10.3390/toxins14120817. PubMed PMID: 36548714. PubMed PMID: PMCPMC9787415.

30. Malhotra A, Wüster W, Owens JB, Hodges CW, Jesudasan A, Ch G, et al. Promoting co-existence between humans and venomous snakes through increasing the herpetological knowledge base. Toxicon X. 2021;12:100081. Epub 20210826. doi: 10.1016/j.toxcx.2021.100081. PubMed PMID: 34522881. PubMed PMID: PMCPMC8426276.

31. Agras S, Sylvester D, Oliveau D. The epidemiology of common fears and phobia. Comprehensive Psychiatry. 1969;10(2):151–6. doi: 10.1016/0010-440X(69)90022-4.

32. Stenson AF, Nugent NR, van Rooij SJH, Minton ST, Compton AB, Hinrichs R, et al. Puberty drives fear learning during adolescence. Dev Sci. 2021;24(1):e13000. Epub 20200728. doi: 10.1111/desc.13000. PubMed PMID: 32497415. PubMed PMID: PMCPMC9206403.

33. Andersson G, Waara J, Jonsson U, Malmaeus F, Carlbring P, Ost LG. Internet-based exposure treatment versus one-session exposure treatment of snake phobia: a randomized controlled trial. Cogn Behav Ther. 2013;42(4):284–91. doi: 10.1080/16506073.2013.844202. PubMed PMID: 24245707.

34. Hunt M, Bylsma L, Brock J, Fenton M, Goldberg A, Miller R, et al. The role of imagery in the maintenance and treatment of snake fear. J Behav Ther Exp Psychiatry. 2006;37(4):283-98. Epub 20060213. doi: 10.1016/j.jbtep.2005.12.002. PubMed PMID: 16473325.

35. McNally RJ. Preparedness and phobias: a review. Psychol Bull. 1987;101(2):283-303. PubMed PMID: 3562708.

36. Rachman S. The conditioning theory of fearacquisition: A critical examination. Behaviour Research and Therapy. 1977;15(5):375–87. doi: 10.1016/0005-7967(77)90041-9.

37. Bandura A. Social learning theory. Englewood Cliffs, NJ: Prentice Hall. 1977.

38. Isbell LA. Snakes as agents of evolutionary change in primate brains. J Hum Evol. 2006;51(1):1-35. Epub 20060320. doi: 10.1016/j.jhevol.2005.12.012. PubMed PMID: 16545427.

39. Kawai N, Koda H. Japanese monkeys (Macaca fuscata) quickly detect snakes but not spiders: Evolutionary origins of fear-relevant animals. J Comp Psychol. 2016;130(3):299-303. Epub 20160414. doi: 10.1037/com0000032. PubMed PMID: 27078076.

40. Sancassiani F, Romano F, Balestrieri M, Caraci F, Di Sciascio G, Drago F, et al. The Prevalence of Specific Phobia by Age in an Italian Nationwide Survey: How Much Does it Affect the Quality of Life? Clin Pract Epidemiol Ment Health. 2019;15:30-7. Epub 20190220. doi: 10.2174/1745017901915010030. PubMed PMID: 30972140. PubMed PMID: PMCPMC6407652.

41. Zsidó AN, Inhof O, Kiss BL, Bali C, March DS. Threatening stimuli have differential effects on movement preparation and execution—A study on snake fear. People and Nature. n/a(n/a). doi: 10.1002/pan3.10500.

42. Pandey DP, Subedi Pandey G, Devkota K, Goode M. Public perceptions of snakes and snakebite management: implications for conservation and human health in southern Nepal. J Ethnobiol Ethnomed. 2016;12(1):22. Epub 20160602. doi: 10.1186/s13002-016-0092-0. PubMed PMID: 27255454. PubMed PMID: PMCPMC4891849.

